# PREDICTORS OF MORTALITY AND CLINICAL CHARACTERISTICS AMONG CARBAPENEM-RESISTANT OR CARBAPENEMASE-PRODUCING ENTEROBACTERIACEAE BLOODSTREAM INFECTION IN SPANISH CHILDREN

**DOI:** 10.1101/2020.05.14.20097188

**Authors:** María Fátima Ara-Montojo, Luis Escosa-García, Marina Alguacil, Nieves Seara, Carlos Zozaya, Diego Plaza, Cristina Schuffelmann-Gutiérrez, Ángela de la Vega, Carlota Fernández-Camblor, Esther Ramos-Boluda, María Pilar Romero-Gómez, Guillermo Ruiz-Carrascoso, Itsaso Losantos-García, María José Mellado-Peña, Rosa Gómez-Gil

**Author notes:** María Fátima Ara-Montojo and Luis Escosa-García contributed equally to this article and both should be considered first author. **María Fátima Ara-Montojo**. Servicio de pediatría hospitalaria, enfermedades infecciosas y tropicales. Hospital Universitario La Paz. Paseo de la Castellana 261, C.P. 28046. Madrid, Spain., **Luis Escosa-García**. Servicio de pediatría hospitalaria, enfermedades infecciosas y tropicales. Hospital Universitario La Paz. Paseo de la Castellana 261, C.P. 28046. Madrid, Spain.

## Abstract

**Background:** Carbapenem-resistant Enterobacteriaceae (CRE) are a growing problem in pediatric population worldwide with high mortality rates (18.5-52%) in bloodstream infection (BSI).

**Objectives:** The aim of this study is to evaluate predictors of 30-day mortality in CRE BSI in a pediatric cohort.

**Methods:** Retrospective observational single-center study (December 2005 - August 2018) was conducted. CRE BSI in children 0 to 16 years were included. Microbiological identification (MALDI Biotyper) and antimicrobial susceptibility testing (Vitek2® and MicroScan panel NBC44) according to current EUCAST breakpoints were performed. PCR OXVIKP® was used to confirm carbapenemases genes (OXA-48, VIM, KPC, NDM). Demographic characteristics, underlying diseases, source of bacteremia, antimicrobial therapy and outcomes were collected from medical records. Survival analysis to establish predictors of 30 day-mortality was performed.

**Results:** Thirty-eight cases were included, 76.3% hospital-acquired infections and 23.7% related to healthcare. All patients had underlying comorbidity and 52.6% had received a transplant. VIM-carbapenemase was the predominant mechanism (92%). Previous CRE colonization or infection rate was 52.6%. Gut (26%) and vascular catheter (21%) were the predominant sources of infection. Crude mortality within 30 days was 18.4% (7/38); directly related 30-day mortality was 10.5%. Conditions associated with an increment in 30-day mortality were intensive care admission and inadequate empiric therapy (p<0.05). Combination antibiotic targeted treatment and a low meropenem MIC were not related to improved survival.

**Conclusions:** CRE BSI mortality rate is high. The most important factor related to 30-day survival in our CRE BSI cohort in children was success in empiric treatment with at least one active antibiotic.

## Introduction

Carbapenem-resistant Enterobacteriaceae (CRE) infections are a growing problem in pediatric population worldwide. In recent years, an increment in the number of cases has been reported in many countries.^1-2^ Some risk factors have been identified in previous studies, including underlying comorbidity, immunosuppression, previous central venous catheter, endotracheal intubation and prior antibiotic exposure.^3,4^

CRE infections in children have been associated with high mortality rates, up to 52%, probably due to delays in appropriate antibiotic therapy, lack of available active antibiotics against CRE, underlying conditions and illness severity. ^5^ Therefore, treatment of CRE infections is a real challenge for clinicians and pediatric antibiotic pipeline is limited. Some studies in adults suggested that carbapenem-containing combination therapy was associated with improved survival in severe invasive infections due to CRE. ^6,7^ Due to the lack of studies in children, data to guide management has been extrapolated from adult literature. Current recommendation in pediatrics is combination therapy of at least two agents with in vitro activity in most scenarios, until more available data and experience with newer betalactam agents.^8-10^ To improve management of CRE infections in pediatric population it is necessary to evaluate if combination therapy is associated with lower mortality rates than monotherapy for all scenarios.

The aim of this study is to evaluate risk factors for CRE bacteremia and independent predictors of mortality (up to 30 days) related to the infection in a pediatric cohort.

## Patients and Methods

### Study design

A retrospective observational single-center study was conducted.

### Setting

Hospital Universitario La Paz (HULP) Children’s Hospital is a tertiary hospital with 256 hospitalization beds belonging to the Spanish Health National System, serving more than 45,000 children’s emergency and more than 8,000 children’s hospital admissions per year in Northern Madrid (Spain). It has a 16-bed pediatric intensive care unit (PICU) and a 23-bed neonatal intensive care unit (NICU), being the National Referral Hospital for pediatric transplantation.

### Inclusion and exclusion criteria

Episodes of bloodstream infection (BSI) in children 0 to 16 years of age that showed the growth of CRE were included from December 2005 to August 2018. CRE isolate was defined as any Enterobacteriaceae non-susceptible to meropenem (MIC > 8 mg/L), imipenem (MIC > 2 mg/L) or ertapenem (MIC > 0.5 mg/L), or if carbapenemase production was documented. ^11^ Polymicrobial bacteremia episodes were excluded.

### Variables and Data Collection

We collected demographic characteristics, reason for admission and diagnosis, underlying diseases, source of bacteremia, empirical and targeted antimicrobial therapy and outcomes from medical records. Hospital’s Microbiology department provided microbiological data.

### Definitions

BSI was classified as hospital-acquired, community-acquired or healthcare-associated (HCA) according to classic and modified CDC criteria. Episodes of community-acquired bacteremia were further classified as healthcare-associated (HCA) if any of the following criteria were present:^12^ 48-hours hospital admission during the previous 90 days, receipt of hemodialysis, intravenous medication or home wound care in the previous 30 days or residence in a nursing home or long-term care facility.

Classification as sepsis or septic shock was performed according to definitions of the International Consensus Conference on Pediatric Sepsis. ^13^

The primary source of bacteremia was determined by the clinical presentation and/or by the evidence of an identical strain cultured near to or on the same date as the onset of BSI from other body site. To classify an infection as catheter-related BSI the 2009 Infectious Diseases Society of America (IDSA) Guidelines criteria were used. ^14^ When the source of bacteremia could not be identified, it was classified as primary bacteremia.

According to Hsu AJ et al. recommendations, antimicrobial therapy was considered “microbiologically appropriate” if the patient received one active agent against the isolate (MIC within the susceptible range) and “clinically adequate” if the patient received a combination of two active antibiotics at a right dose and route according to source of infection. ^8^ Use of imipenem or meropenem (MIC ≤ 8 mcg/ml) was considered adequate if high-dosed (25 mg/kg q6h for imipenem, 40 mg/kg over 3 hours q8h for meropenem), adjusted to renal function, and associated with a second microbiologically active agent.

### Bacterial isolates and microbiological work-up

Blood cultures were incubated in the Bactec automated blood culture device (BACTEC ^TM^, Becton Dickinson, Franklin Lakes, NJ, USA) and BacT/ALERT® (bioMérieux, Marcy l’Etoile, France) blood culture bottle systems. All positive blood cultures were routinely subcultivated on three agar plates: sheep blood agar, chocolate blood agar and *Brucella* blood agar.

Identification was performed by MALDI Biotyper (Bruker Daltonik GmbH, Bremen, Germany) and antimicrobial susceptibility testing (AST) was performed using the automated system Vitek2®, (bioMérieux Marcy l’Etoile, France) and MicroScan panel NBC44 (Beckman Coulter) according to European Committee on Antimicrobial Susceptibility Testing (EUCAST) breakpoint. ^15^ Isolates were tested for extended spectrum beta-lactamases (ESBL) production using E-test ESBL stripes (bioMérieux) and the double-disk synergy test with aztreonam and amoxicillin/clavulanic acid. Isolates having a MIC > 1 mg/L to imipenem or meropenem and > 0.5 mg/L to ertapenem were confirmed by PCR (OXA-48, VIM, KPC, NDM specific primers) with OXVIKPND® real time PCR kit (Progenie Molecular, Spain).

Genetic relationships between isolates were determined by multilocus-sequence typing and automated repetitive-sequence-based PCR in *Klebsiella pneumoniae* using the DiversiLab® (bioMérieux) system.

### Outcome parameters

Primary outcome was death within 30 days after the first positive CRE blood culture. Relationship between death and BSI episode (directly related or not directly related) was established by two independent investigators, by reviewing hospital medical records.

### Statistical analysis

Categorical variables were presented as absolute frequencies and percentages. Continuous variables were presented as the mean or median, with its standard deviation or ranges, respectively. Categorical variables were compared using chi-square test or Fisher’s Exact test, as appropriate. Continuous variables were compared by Student’s t test or Mann-Whitney U test according to their distribution. Univariate analysis was conducted to evaluate different possible factors related to mortality. The survival analysis was carried out by means of the Kaplan-Meier analysis and “log-rank test” was performed to compare survival functions. A two-tailed *P* value < 0.05 was considered statistically significant. All statistical analysis was performed using SAS 9.3 (SAS Institute, Cary, NC, USA).

### Ethics

HULP Institutional Review Board evaluated and approved the study protocol. All collected data were anonymized. Given its observational retrospective design, written informed consent was waived.

## Results

### Clinical and epidemiological characteristics

Of 2842 positive blood cultures in children during the study period, 928 (32.7%) were Enterobacteriaceae and 38 (1.3%) were CRE isolates that met inclusion criteria (Table 1). Most cases were detected between 2010 and 2015 (73.6%). Mean age was 2.2 years (DS 3.2, range 0-14.5) and patients were predominantly female (55.3%).

**TABLE 1.**
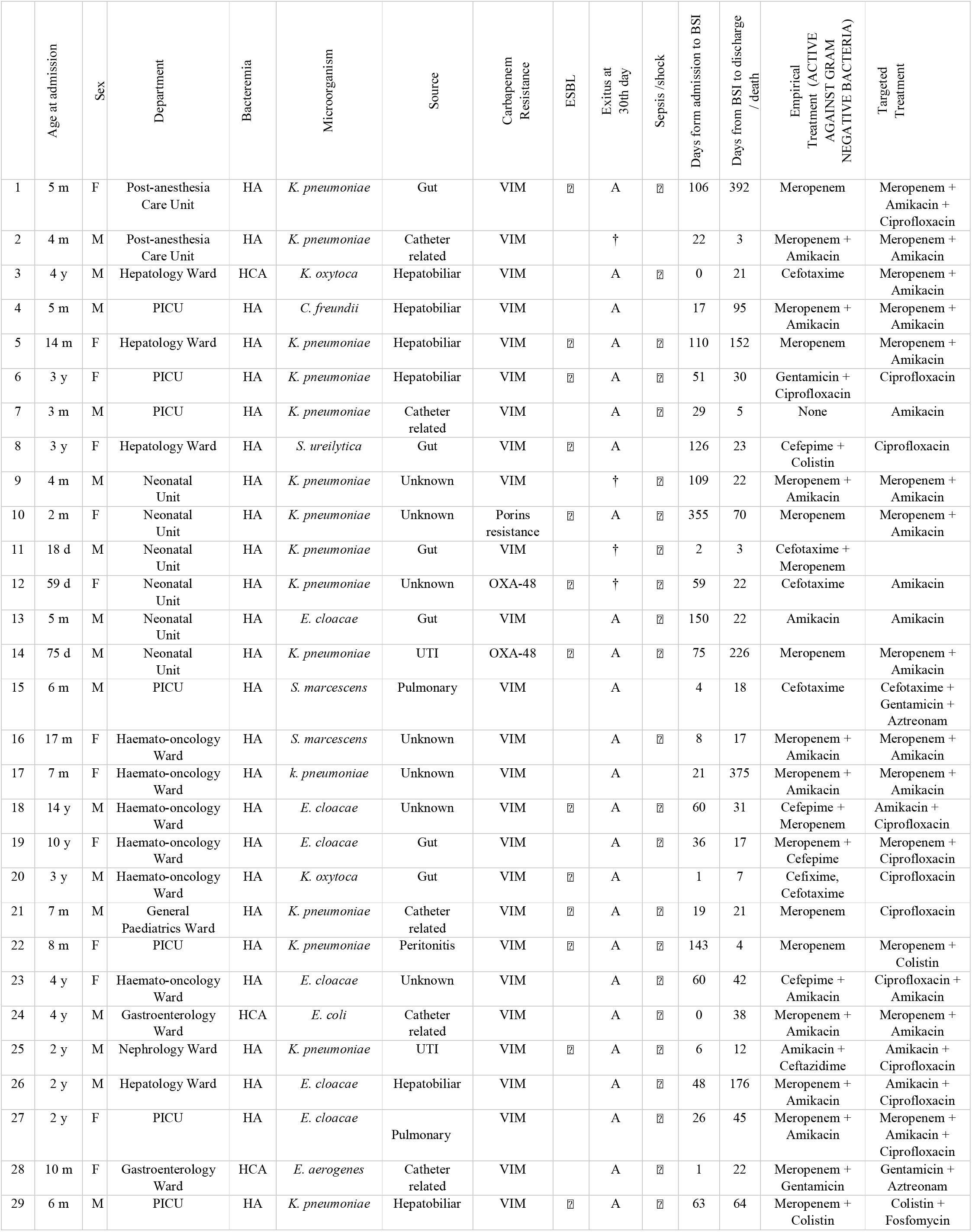
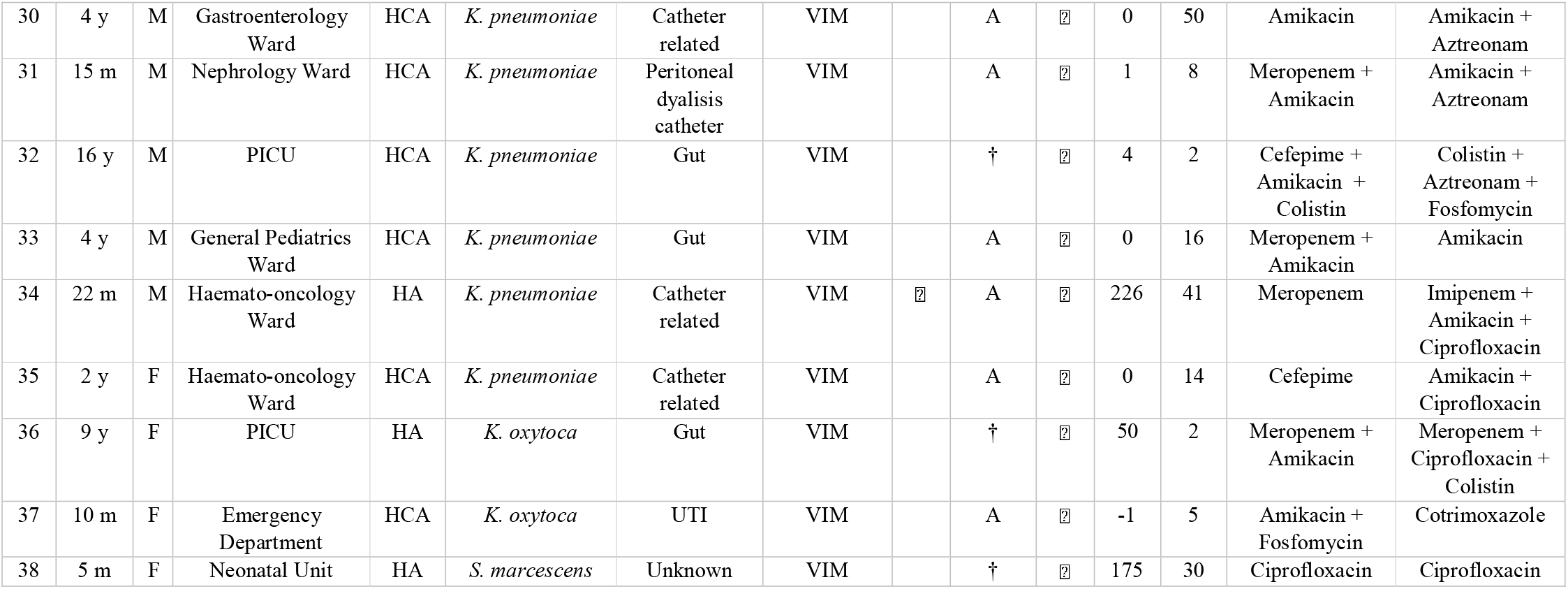
Clinical features of patients with bacteremia due to CRE. Months (m), female (F), hospital-acquired (HA), alive (A), exitus (+), male (M), years (y), health care associated (HCA), days (d), urinary tract infection (UTI).

No community-acquired cases were detected. Twenty-nine (76.3%) cases were hospital-acquired infections and 9 (23.7%) were related to healthcare.

In regard to underlying conditions, 20 patients (52.6%) had received a transplant. Solid organ transplant was documented in 13 (9 liver, 3 multivisceral and 1 kidney), an allogenic hematopoietic stem cell transplantation in 6 patients and 1 patient had received both (multivisceral and allogeneic hematopoietic stem cell transplantation). Other medical conditions were: heart disease (5/38, 13.2%), neurological (4/38; 10.5%), respiratory (4/38, 10.5%), urological (3/38, 7.9%), digestive (3/38, 7.9%) and malignancy (one hematologic and one solid organ tumor). Fourteen (36.8%) patients were born prematurely.

At the time of bacteremia, 51.7% of patients with hospital-acquired disease were hospitalized in intensive care units (8 in PICU, 7 in NICU), 41.4% in a medical ward and 6.9% were in a surgical ward (Post-Anesthesia Care Unit).

The rate of previous antibiotic exposure, indwelling devices and other possible risk factors for CRE infection are shown in Table 2.

**TABLE 2.**
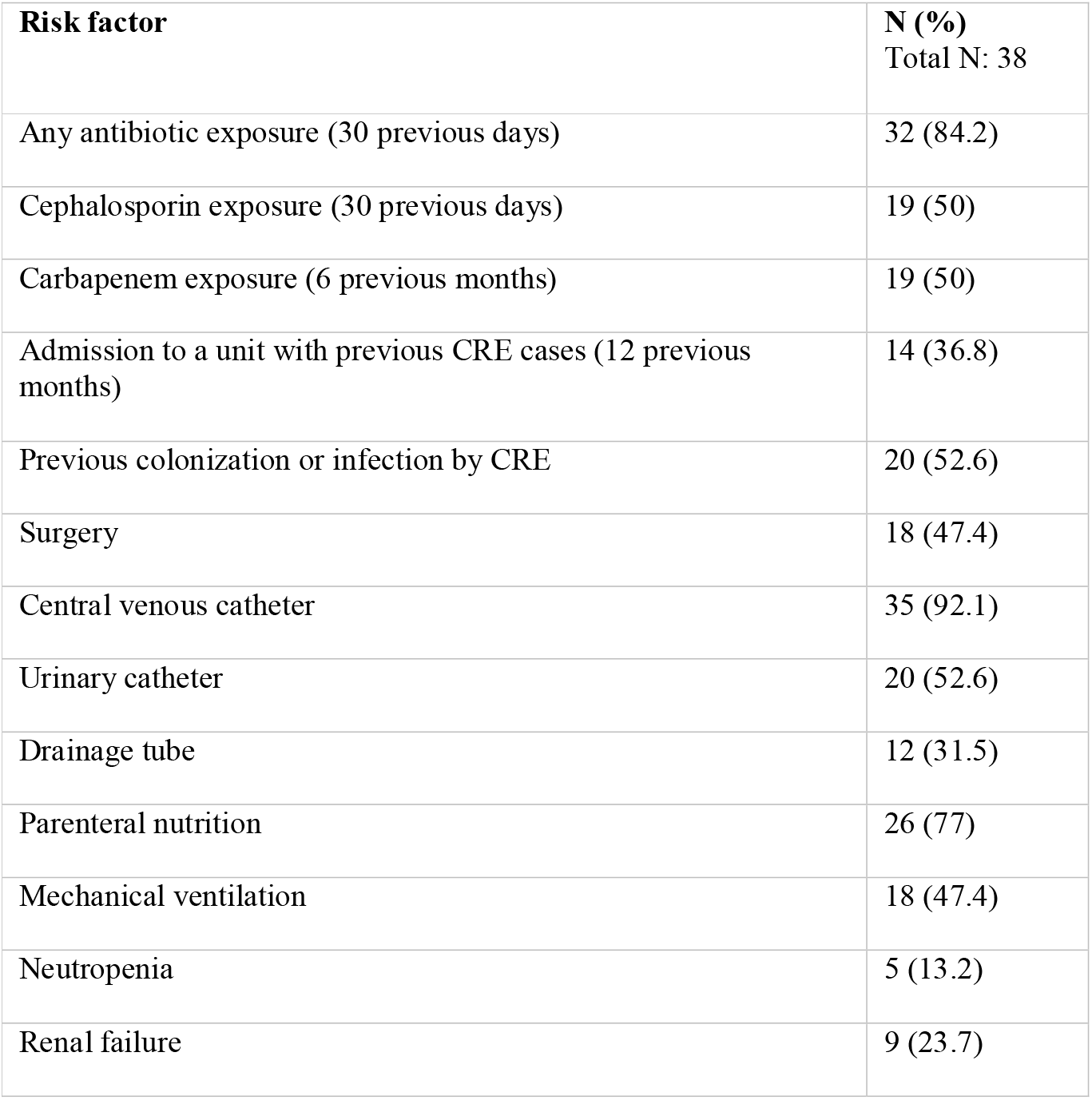
Risk factors related to CRE bacteremia.

Regarding the source of bacteremia, the intestinal tract was found to be the most common origin causing 11 cases (26.3%), followed by central venous catheter-related bacteremia in 8 (21.1%) and hepatobiliary origin in 6 (5.8%) patients. Three (7.9%) children had urinary tract infection and one patient had a peritoneal catheter-related infection. Only one patient had ventilator-associated pneumonia. The origin remained unknown in 8 (21.1%) cases.

As per severity of disease, 32 (84.2%) patients with CRE BSI met systemic inflammatory response syndrome (SIRS) criteria of which 8 (21.1%) presented as septic shock.

### Microbiological results

The isolated bacteria, resistance mechanisms and susceptibility to antimicrobials are shown in Table 3.

**TABLE 3.** Resistance of strains to antimicrobials agents. “MIC” is the Minimum Inhibitory Concentration acquired in mg/L; “Range MIC” is the range of Minimum Inhibitory Concentration; “S” stands for susceptible; “I” for intermediate and “R” for resistant, as interpreted according to the EUCAST V 6.0 guidelines. “AZT” for aztreonam, “ERT” for ertapenem, “IMI” for imipenem, “MER” for meropenem, “COL” for colistin, “AK” for amikacin”, “CIP” for ciprofloxacin, “TYG” for tygecicline, “FOS” for fosfomycin.

| Strain |  |  |  |  |  |  |  |  |  |  |
| --- | --- | --- | --- | --- | --- | --- | --- | --- | --- | --- |
|  |  | AZT | ERT | IMI | MER | COL | AK | CIP | TYG | FOS |
| *K. pneumoniae* VIM (18) | Range MIC | ≤1 - >8 | 1 - >1 | 2 - >8 | ≤1 - >8 | ≤0.5->4 | ≤2 ->32 | ≤0,5 ->2 | ≤1 - >4 | ≤32->64 |
|  | S (%) | 50 | 0 | 11.1 | 44.4 | 83.3 | 72.2 | 61.1 | 77.7 | 83.3 |
| *K. pneumoniae* OXA-48 ESBL (2) | Range MIC | >16 | >1 | 2-8 | 2-8 | ≤0.5 | ≤2 | >2 | 4 | >64 |
|  | S (%) | 0 | 0 | 50 | 50 | 100 | 100 | 0 | 0 | 0 |
| *K. pneumoniae* ESBL (1) | MIC | >16 | >1 | 8 | 8 | ≤0.5 | ≤2 | >2 | ≤1 | ≤16 |
|  | S (%) | 0 | 0 | 0 | 0 | 100 | 100 | 0 | 100 | 100 |
| *E. cloacae* VIM (6) | Range MIC | ≤1 - >8 | ≤0.5->1 | 4 - >8 | ≤1 - >8 | ≤0.5 ->4 | ≤2 | ≤0.5 - 1 | ≤1 | ≤16 |
|  | S (%) | 66.6 | 50 | 16.6 | 50 | 66.6 | 100 | 66.6 | 100 | 83.3 |
| *K. oxytoca* VIM (4) | Range MIC | ≤1->32 | >32 | ≤1->8 | ≤1->8 | ≤0.5 | ≤2->32 | ≤0,5 | ≤1 | ≤32->64 |
|  | S (%) | 50 | 0 | 25 | 50 | 100 | 75 | 100 | 100 | 25 |
| *S. marcescens* VIM (3) | Range MIC | ≤1 | >1 | >8 | >8 | >2 | ≤2 | ≤0,5 | ≤1 | ≤32->64 |
|  | S (%) | 100 | 0 | 0 | 0 | 0 | 100 | 100 | 66.6 | 66.6 |
| *E.coli* VIM (1) | MIC | ≤1 | 1 | 8 | 8 | ≤0.5 | ≤2 | ≤0.5 | ≤1 | 32 |
|  | S I R | S | I | I | I | S | S | S | S | S |
| *C. freundii* VIM (1) | MIC | ≤1 | >1 | >8 | >8 | ≤0.5 | ≤2 | ≤0.5 | ≤1 | ≤16 |
|  | S I R | S | R | R | R | S | S | S | S | S |
| *S. ureilytica* VIM (1) | MIC | >8 | 1 | >8 | 2 | >4 | ≤2 | ≤0,5 | ≤1 | 32 |
|  | S I R | R | I | R | S | R | S | S | S | S |
| *E. aerogenesa* VIM (1) | MIC | >8 | 1 | >8 | 2 | >4 | ≤2 | ≤0,5 | ≤1 | 32 |
|  | S I R | R | I | R | S | R | S | S | S | S |
| ALL (38) | S (%) | 55.3 | 7.9 | 13.2 | 39.5 | 73.6 | 84.2 | 68.4 | 81.6 | 71.1 |
|  | I (%) | 2.6 | 7.9 | 36.8 | 34.2 | --- | --- | --- | 7.9 | --- |
|  | R (%) | 42.1 | 84.2 | 50 | 26.3 | 26.3 | 15.8 | 31.6 | 10.5 | 28.9 |

### Treatment and outcomes

Crude mortality within 30 days from onset of BSI was 18.4% (7/38). Death was considered directly related to the BSI episode in 10.5% of total cases, based on clinical judgment.

Before antibiotic susceptibility test results, 10 (26.3%) children received microbiologically inappropriate empiric therapy (no active antibiotic) while the remaining 28 (73.7%) patients were treated with at least one active antibiotic. In 23 (60.5%) cases a carbapenem was used before knowing antibiotic susceptibilities. Median duration of empiric treatment was 3 days (IQR 2-5). Optimal early source control was performed in 23.7% (9/38).

Thirty-six (94.7%) patients received microbiologically appropriate targeted therapy and 15 of them (39.5%) were clinically adequately treated with at least two active antibiotics, at a proper dosage and route according to the source of infection.

In univariate analysis, we found a significantly higher mortality rate in patients admitted to neonatal and intensive care units at the time of BSI onset. Including one active antibiotic in the empiric therapy showed to decrease mortality risk in a 92.4% compared to non-active antibiotic (HR 0.076; IC 95% 0.012-0.494). There was no difference in survival depending on meropenem MIC and neither between patients who received meropenem as empiric therapy and those who not. Attending targeted treatment, we did not find differences when monotherapy was used versus combination therapy and the use of meropenem did not prove to improve survival either (Table 4, Figure 1).

**TABLE 4.** Univariate analysis of risk factor associated with 30-day mortality. The survival analysis was carried out by means of the Kaplan-Meier analysis and “log-rank test” was performed to compare the survival functions. A two-tailed P value < 0.05 was considered statistically significant. Non-significant (NS). * One patient excluded because of death before knowing antibiogram.

| **Variable** | **Survived**<br>**N=31 (%)** | **Died**<br>**N=7 (%)** | **Hazard ratio**<br>**(95% CI)** | **P** |
| --- | --- | --- | --- | --- |
| Median Age (months) | 17.64 | 4.32 |  |  |
| Male (%) | 54.84 | 57.14 |  |  |
| Klebsiella isolated | 19 (61.29) | 6 (85.71) |  | NS |
| Source of infection: |  |  |  |  |
| Intestinal or hepatobiliary tracts | 13 (41.94) | 3 (42.86) |  | NS |
| Catheter-related | 7 (22.58) | 1 (14.29) |  | NS |
| Urinary tract | 3 (9.68) | 0 (0) |  | NS |
| Pulmonary | 2 (6.45) | 0 (0) |  | NS |
| Peritoneal catheter | 1 (3.23) | 0 (0) |  | NS |
| Unknown | 5 (16.13) | 3 (42.86) |  | NS |
| Neonatal unit admission | 3 (9.68) | 4 (57.14) | 0.197 (0.111-0.956) | **0.017** |
| Intensive care unit admission | 10 (32.26) | 6 (85.71) | 0.13 (0-0.984) | **0.024** |
| Presentation as sepsis | 26 (83.87) | 6 (85.71) |  | NS |
| Presentation as septic shock | 5 (16.13) | 3 (42.86) |  | NS |
| Meropenem MIC >8 mg/L | 8 (25.81) | 2 (28.57) |  | NS |
| Optimal early source control | 8 (25.81) | 1 (14.29) |  | NS |
| No-active antibiotic in empiric therapy | 6 (19.35) | 4 (57.14) | 0.076 (0.012-0.494) | **0.001** |
| Meropenem in empiric therapy | 19 (61.29) | 4 (57.14) |  | NS |
| Aminoglycoside in empiric therapy | 15 (48.39) | 4 (57.14) |  | NS |
| Non-clinically adequately or improvable target therapy | 19 (61.29) | 3/6 (50)* |  | NS |
| Monotherapy in target treatment | 16 (51.61) | 3/6 (50)* |  | NS |
| Meropenem in target therapy | 14 (45.16) | 4/6 (66.66)* |  | NS |

**FIGURE 1.**
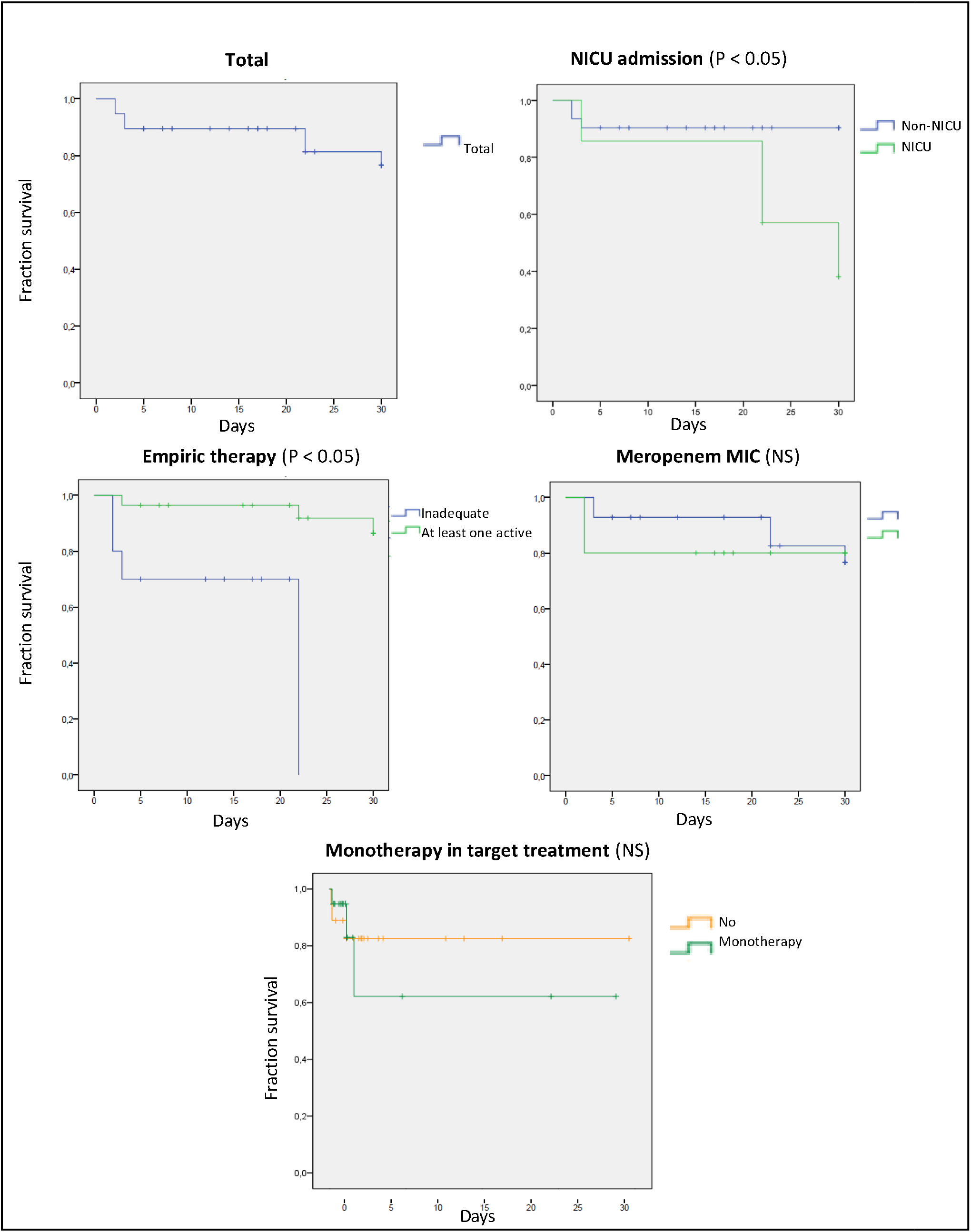
Kaplan-Meier curves of different factors and survival at 30 days in patients with CRE bacteremia. Non-significant (NS); Minimum Inhibitory Concentration (MIC)

## Discussion

In the present study, we report 38 cases of CRE bacteremia in pediatric population. To the best of our knowledge, this is the largest current pediatric cohort reported in Europe. Novel agents (ceftazidime-avibactam, aztreonam-avibactam, cefiderocol, etc.) were not available during the study period, so only classical therapeutic agents were used. VIM-type metallo-beta-lactamases were the predominant resistance mechanism in our series, as they were present in 35 of the 38 strains. These VIM-type enzymes were first discovered in Europe in the 1990s and have been reported worldwide in the recent multidrug-resistant Enterobacteriaceae crisis, although the Mediterranean basin and the Middle East are the predominant areas. ^16^ Most of our cases (73.6%) were detected between 2010 and 2015, due to a VIM 1 *Klebsiella pneumoniae ST-54* outbreak during 2010 and an OXA-48 *Klebsiella pneumoniae ST-11* outbreak in 2012. Antimicrobial stewardship program initiatives in children emerged in 2014 in our hospital, primarily focused on PICU patients so we only found one CRE BSI after 2015.

All episodes of infection in our study were either hospital-acquired or associated with healthcare as in other similar series. ^3,17^ Regarding risk factors of CRE BSI, we found that those children who developed CRE BSI had multiple comorbidities, most of them (52.6%) had received a transplant since our hospital is National Referral Hospital for pediatric transplantation. Of note, 94.7% of patients were carriers of indwelling devices. In addition, they often had exposure to carbapenems in the previous 6 months (50%).

The 30-day mortality rate in our study (18.4%) is not as high as compared with previous reports, ranged from 18.5 to 52%. ^5,18-20^ Grouping data from those reports and our cohort, mortality rate is 31.8% in pediatric CRE bacteremia, although important differences between series are obvious, including different resistance mechanisms or bacteremia severity. For instance, our lower mortality rate could be related to the relatively low rate of severe bacteremia in our series (21.1% had septic shock) and also to the fact that many strains were susceptible to aminoglycosides (81.6%), offering therapeutic options. In India, Nabarro et al. found a mortality rate of 52%, but resistance to aminoglycosides in their cohort was up to 90%. ^5^ In a recent study in Chinese pediatric population with carbapenem-resistant *K.pneumoniae* BSI, mortality rate was similar than ours in relation to low clinical severity and hematological malignancies predominance whose empirical therapy used to be more effective in covering multidrug-resistant bacteria. ^18^ In a meta-analysis of carbapenem-resistant *K. pneumoniae* infections, mortality was not shown to be lower in VIM-producing compared to KPC. Therefore it is unlikely that the VIM predominant resistance mechanism in our cohort explains our lower mortality rate.^21^

We have found a relationship between increased 30 day-mortality and inadequate empirical antibiotic therapy (no active agent), as was shown in previous studies. ^22^ A recent report by Lodise et al. has established that delayed appropriate therapy may be a more important driver of outcomes than CRE itself. ^23^ However, Zhang et al. did not identify an association between non-active empirical antibiotic agents and mortality in children. ^18^ Of note, 52.6% of patients in our cohort had been colonized or infected by a CRE before, as systematic CRE colonization screening in our hospital is performed since 2010 in many units. In adults, Girmenia et al. showed a rate of CRE infection in previously colonized-patients of 10% for immunocompetent, 25% for those with neutropenia, 26% for auto-SCT (stem cells transplant) and 39% for allo-SCT. ^24^ In our pediatric hospital, an infection rate of 13% has been established in patients with previous VIM-type CRE colonization ^25^, but from our knowledge no pediatric data has set which colonized children are at a higher risk to develop an infection. As many patients in our cohort were previously CRE colonized and active antibiotic in empiric therapy showed to decrease mortality risk in a 92.4% compared to non-active antibiotic, antimicrobial stewardship guided antibiogram in colonized stools from selected highly-risk patients might be an interesting strategy to improve CRE BSI prognosis. However, rapid diagnostic testing aiming to reduce the time to effective therapy in antimicrobial stewardship programs would be the most desirable approach. ^26^

Patients admitted to neonatal units and to intensive care units also had significant higher mortality rates in our cohort. Of note, in our survival analysis, there was no evidence of higher mortality in patients with isolated *Klebsiella* spp. strains. No source of infection showed relation to mortality either. Relationship between meropenem MIC and increased mortality has previously been described in adults and Nabarro and Zhang also found statistically increased mortality rate in pediatric patients with CRE BSI with a meropenem MIC > 8mg/L. ^5,18^ Nevertheless, in our cohort meropenem MIC > 8 was not predictive of higher mortality. Empiric antibiotic therapy that included meropenem was also no protective in Zhang’s et al. report. Regarding the use of aminoglycosides as empiric treatment, we did not found differences in mortality either.

In adults, retrospective observational studies have suggested improved survival in patients receiving combinations of two or more in vitro active antibiotics, mostly among patients with a high probability of death. ^6^ These data have been extrapolated to children with a number of authors suggesting use of combination antibiotics in CRE. ^8^ The few pediatric series so far show controversy in this aspect. In our study, targeted treatment with combinations of two or more active agents did not lead to higher survival than monotherapy, as has been shown in a recent clinical trial in adults, where combination therapy was not superior to monotherapy. ^27^ However, most infections in Paul et al. trial (77%) were caused by *Acinetobacter baumanii* so CRE might have been under-represented. The inclusion of meropenem in the targeted therapy also showed no survival improvement in our cohort.

Our study has some limitations. It is a retrospective study of a single cohort from just one center so the sample size is limited, although it is the largest pediatric series reported in Europe so far. This may have influenced the statistical power to identify risk factors and predictors of mortality. Moreover, the high proportion of VIM-producing Enterobacteriaceae in our cohort leads to results that may not be representative for all CRE. Regarding surveillance analysis, our low mortality rate decreases the reliability of results and we have not used severity scores. Nevertheless, given the scarcity of literature on this important subject, mostly in Western Europe, we believe it offers useful insights into the management of CRE BSI in children.

The findings of this study suggest CRE BSI are a growing problem in pediatric patients with comorbidities and underlying devices. The independent factor more related to 30-day mortality in our cohort was success in empiric treatment with at least one active antibiotic. Antimicrobial stewardship strategies focusing on the adequacy of empiric therapy are needed in CRE BSI. Combination antibiotic targeted treatment and a low Meropenem MIC were not related to improved survival in our cohort but sample size might limit these findings.

## Data Availability

We collected patients data from medical records in an anonymized Excel database.

## Transparency declarations

María Fátima Ara-Montojo and Luis Escosa-García contributed equally to this article and both should be considered first author. We have nothing to declare.

## Transparency declarations

None to declare.

